# Comparative study of the treatment of mechanical jaundice in men and women: a cross-sectional study

**DOI:** 10.1101/2022.12.28.22284006

**Authors:** Basheer Abdullah Marzoog, Kostin Sergey Vladimirovich

**Affiliations:** National Research Mordovia State University

**Keywords:** Cholithiasis, Pathophysiology, Surgery, Obstructive Jaundice, Antibiotic: Pathogenesis

## Abstract

**Background:** Cholithiasis remains the leading cause of obstructive jaundice. A tendency to cholithiasis is suggested in women. However, the underlying risk factors and statistical conformation are lacking.

**Aims:** Retrospectively describe and assess the causes of obstructive jaundice, as well as demonstrate the changes in laboratory parameters in response to treatment.

**Objectives:** The study describes a sample of patients with obstructive jaundice due to various causes.

**Material and methods:** A retrospective cohort study involved 101 patients with cholithiasis for the period 14.01.2016-13.04.2018 treated surgically and or conservatively. The study involved 60 (59.40594 %) men and 41 (40.59406 %) females aged 16 to 100 years (mean; 64.9901, Std Err: 1.53787). Of 101, 54 (53.46535 %) patients live in the city and 47 (46.53465 %) live in the village. The patients passed a full blood count and biochemical analysis for at least two times. Data were collected from the Mordovian Republic Hospital and retrospectively analyzed. The consent of the patients has been taken for scientific purposes to analyze and publish the results of the study. For statistical analysis, used T test, one way ANOVA test, and Spearman correlation test by using Statistica program.

**Results:** By the etiology of obstructive symptoms, the frequency of gallstones is reported in 37 (36.63366%) patients, acute pancreatitis in 23 (22.77228%) patients, post-cholecystectomy syndrome (PCS) in 8 (7.92079 %) patients, Hilar cholangiocarcinoma (HC, Klatskin tumor) in 1 (0.99010 %) patient, pancreatic cancer in 12 (11.88119 %) patients, acute biliary pancreatitis in 8 (7.92079 %) patients, pancreatic pseudotumor in 4 (3.96040%) patients, acute cholecystitis in 3 (2.97030 %) patients, papillary tumor in 2 (1.98020 %) patients, and pancreatic cyst in 3 (2.97030%) patients. In male group, the mean age is 62.7805 years (min; max, 16.00000; 86.000) years, (median; Std Err, 66.0000; 2.40541). In the female group, the mean age is 66.5000 years (min; max, 24.00000; 100.000) years, (median; Std Err, 65.5000; 1.99300). Of 101 patients, 20 (19.80%) patients underwent surgical treatment and 81 (80.20%) patients did not require surgery. The mean total hospitalization days for patients who passed EPST surgery is 16.20000 days (Std Err 1.008850), CBD 21.50000 days (Std Err 1.565248), CDBD 25.00000 days, cholecystostomy 14.00000 days and hepaticocholecystoenterostomy 16.00000 days (Std Err 2.000000). In male group, the mean total hospitalization days 15.8537 (min; max, 5.00000; 30.000) days, (median; Std Err, 15.0000; 0.89071). In the female group, the mean total hospitalization days 14.0833 (min; max, 6.00000; 29.000) days, (median; Std Err, 13.5000; 0.68901). A direct association between the glucose value and the age, the correlation coefficient value -0.961980.

**Conclusions:** Tendency to the early occurrence of obstructive jaundice symptoms in men compared to women. In treatment plans, men and females required the same total hospitalization days. The incidence rate of cholithiasis in females is higher than in males.

**Other findings:** A straight association between age and the etiology of obstructive jaundice symptoms as well as a straight association between total hospitalization days and the type of surgery.

## Introduction

Gallstone disease is a common medical condition worldwide, with 10–20% of patients with choledocholithiasis developing biliary pancreatitis and cholangitis. A majority of stones form in the gallbladder and then pass into the common bile duct, where they generate symptoms, due to biliary obstruction. The most certain symptomatic manifestation of gallstones is episodic upper abdominal pain. Dyspeptic symptoms of indigestion, belching, bloating, abdominal discomfort, and heartburn [1–5].

Gallbladder stones classified into several types, including cholesterol stones, pigment stones, calcium carbonate stones, phosphate stones, cystine stones and mixed stones [6, 7].

Obstructive jaundice (OJ) is a term that describes the clinical entity of yellowness of the skin and mucous membranes due to the inability of bile to flow freely into the duodenum. This is commonly due to mechanical or physiological blockage of either the intrahepatic or extrahepatic bile ducts [8] [9]. Obstructive jaundice is a common problem in daily clinical practice. Understanding the pathophysiological changes in obstructive jaundice remains a challenge for planning current and future treatment [9, 10].

According to the etiopathogenesis, OJ is fundamentally divided into benign (non-tumor) and malignant (tumor). The main cause of benign OJ in adults is obstruction of the common bile duct or large duodenal papilla with calculus in gallstone disease; while malignant OJ mainly develops from tumor lesions of the pancreatic head, which block the free removal of bile into the duodenum [11–13].

Laboratory diagnostics include her white blood count, C-reactive protein (CRP), aspartate aminotransferase (AST), alanine aminotransferase (ALT), Υ-glutamyl transpeptidase (GGT), alkaline phosphatase (AP), and bilirubin levels [14, 15].

The array of invasive and noninvasive radiological techniques commonly employed for investigating hepatobiliary lesions includes computed tomography (CT), percutaneous transhepatic cholangiography (PTC), endoscopic ultrasound (EUS), endoscopic retrograde cholangiopancreatography (ERCP), helical CT cholangiography, Magnetic Resonance Cholangiopancreatography (MRCP), radionuclide imaging, and ultrasonography [16, 17].

Treatment of common bile duct stones and gallstones includes open biliary exploration, cholecystectomy, laparoscopic cholecystectomy, and endoscopic sphincterotomy (ES)□with□laparoscopic cholecystectomy [18–20].

The majority of gallstone carriers will remain asymptomatic and about one in five will develop symptoms. Determinants of the progression of the disease from asymptomatic to symptomatic disease include sex, age, body mass index, smoking, and gallstone ultrasound characteristics [21–25].

During the reproductive years, the female-to-male ratio is about 4:1, with the sex discrepancy narrowing in the older population to near equality. The risk factors predisposing to gallstone formation include obesity, diabetes mellitus, estrogen and pregnancy, hemolytic diseases, and cirrhosis [26, 27].

The study sought to assess the treatment plan in men and women, including the total days of hospitalization and the surgery used in the treatment of the patients.

## Materials and methods

A retrospective cohort study involved 101 patients with cholithiasis for the period 14.01.2016- 13.04.2018 treated surgically and conservatively. The primary descriptive statistics results demonstrated that a gender dependent potential differences exist and therefore we divided the sample on gender dependent base. The study involved 60 (59.40594 %) men and 41 (40.59406 %) women aged 16 to 100 years (mean; 64.9901, Std Err: 1.53787). Of 101, 54 (53.46535 %) patients live in the city and 47 (46.53465 %) live in the village. The patients passed a full blood count and biochemical analysis. Data were collected from the Mordovian Republic Hospital and retrospectively analyzed. The consent of the patients has been taken for scientific purposes to analyze and publish the results of the study.

For statistical analysis, used T test, one way ANOVA test, and Spearman correlation test by using Statistica program (StatSoft, Inc. (2011). STATISTICA (data analysis software system), version 10. www.statsoft.com.). The results of the described t test are all statistically significant at p<0.05 and the non-statistically significant results are not described. The results first describe the whole sample, and then divide the results into male and female groups. Units of measurement of bilirubin, alanine transaminase (ALT), aspartate transaminase (AST), and glucose are Mmol / L or U / L. The measurement units of leukocytes (WBC) and erythrocytes (RBC) presented by cubic millimeter (million/mm3).

## Results

The descriptive statistics results showed that mean total hospitalization days of the patients is 14.8020, Std Err 0.55025 (median: min; max, 14.0000: 5.00000; 30.000). The mean glucose level is 5.9824, Std Err 0.36773 (median: min; max, 14.0000: 5.00000; 30.000). The mean level of Hb1 is 125.0000, Std Err 2.12205 (median: min; max, 125.0000: 80.00000; 170.000). The mean Hb2 level is 46.3500, Std Err 5.94892 (median: min; max, 11.6500: 4.00000; 156.000). The mean level of WBC1 is 8.2253, Std Err 0.36721 (median: min; max, 7.1000: 3.40000; 18.900). The mean level of WBC2 is 5.6706, Std Err 0.28519 (median: min; max, 5.0000: 3.34000; 14.700). The mean level of RBC1 is 4.0635, Std Err 0.08855 (median: min; max, 4.1050: 2.80000; 5.170). The mean level of RBC2 is 13.1448, Std Err 2.84458 (median: min; max, 4.2000: 3.20000; 44.000). The mean level of Ht1 is 33.6995, Std Err 1.86251 (median: min; max, 35.5000: 0.39000; 43.000). The mean level of Ht2 is 174.5672, Std Err 25.91792 (median: min; max, 98.0000: 22.00000; 1500.000). The mean AST1 level is 98.4900, Std Err 9.67792 (median: min; max, 76.0000: 12.00000; 568.000). The mean AST2 level is 171.9881, Std Err 25.85144 (median: min; max, 101.0000: 4.00000; 1900.000). The mean level of ALT1 is 124.0838, Std Err 12.28858 (median: min; max, 96.5000: 16.00000; 746.000). The mean level of ALT2 is 101.1956, Std Err 8.15750 (median: min; max, 78.5000: 10.00000; 427.000). The mean total level of bilirubin 1 is 118.2385, Std Err 20.39748 (median: min; max, 59.0000: 5.40000; 1645.000). The mean total bilirubin 2 level is 72.5812, Std Err 7.64901 (median: min; max, 48.5000: 0.00000; 451.000). The mean level of direct bilirubin 1 is 59.9176, Std Err 7.33694 (median: min; max, 28.0000: 0.00000; 302.000). The mean level of direct bilirubin 2 is 44.6942, Std Err 4.50862 (median: min; max, 30.0000: 0.00000; 231.000). The mean indirect bilirubin 1 is 47.9066, Std Err 6.43306 (median: min; max, 25.0000: 0.00000; 432.000). The mean level of indirect bilirubin 2 is 18.8347, Std Err 3.41557 (median: min; max, 6.4000: 0.00000; 220.000).

In terms of correlations, we found a straight association between age and WBC2 level, correlation coefficient value -0.973374. Direct association between red blood cells 2 (RBCs) level and total hospitalization days, correlation coefficient value 0.962094. A direct association between the hematocrit 1 (Ht) value and the age, correlation coefficient value -0.968686. A straight association between the ALT1 value and total hospitalization days, correlation coefficient value 0.978209. A direct association between the total bilirubin 1 value and hemoglobin 1 (Hb) and Hb2, correlation coefficient value -0.963832; -0.973739, respectively. A straight association between the indirect bilirubin 1value and the Hb1 and Hb2, correlation coefficient value -0.955833; -0.964377, respectively. A direct association between the glucose value and the age, the correlation coefficient value -0.961980. A straight association between the hematocrit value and the age, correlation coefficient value -0.968686. A straight association between the Ht1 value and the Hb1, Hb2, and WBC1, correlation coefficient value -0.993404, 0.983844, and 0.955230, respectively. A straight association between the RBC2 value and AST1 and ALT1, correlation coefficient value 0.988016, 0.994187 respectively. A straight association between the ALT1 value and AST1, correlation coefficient value 0.965974. A straight association between the ALT2 value and AST2, correlation coefficient value 0.971589. A direct association between the total bilirubin 1 value and AST1, correlation coefficient value 0.964113. A straight association between the total bilirubin 2 value and the AST2, correlation coefficient value 0.959727. A direct association between the direct bilirubin 2 value and the AST2, the correlation coefficient value 0.971398. A straight association between the indirect bilirubin 1 value and AST1, correlation coefficient value 0.952995. A straight association between the glucose value and the value of the Ht 2, correlation coefficient 0.979958. A straight association between the indirect bilirubin 1 value and total bilirubin 1, correlation coefficient value 0.998422. A straight association between the indirect bilirubin 2 value and total bilirubin 2, correlation coefficient value 0.997908. A direct association between the value of direct bilirubin 2 and the value and total bilirubin 2, correlation coefficient 0.997026. A straight association between the indirect bilirubin 2 value and direct bilirubin 2, correlation coefficient value 0.989957. (*Figure 1*)

**Figure 1:**
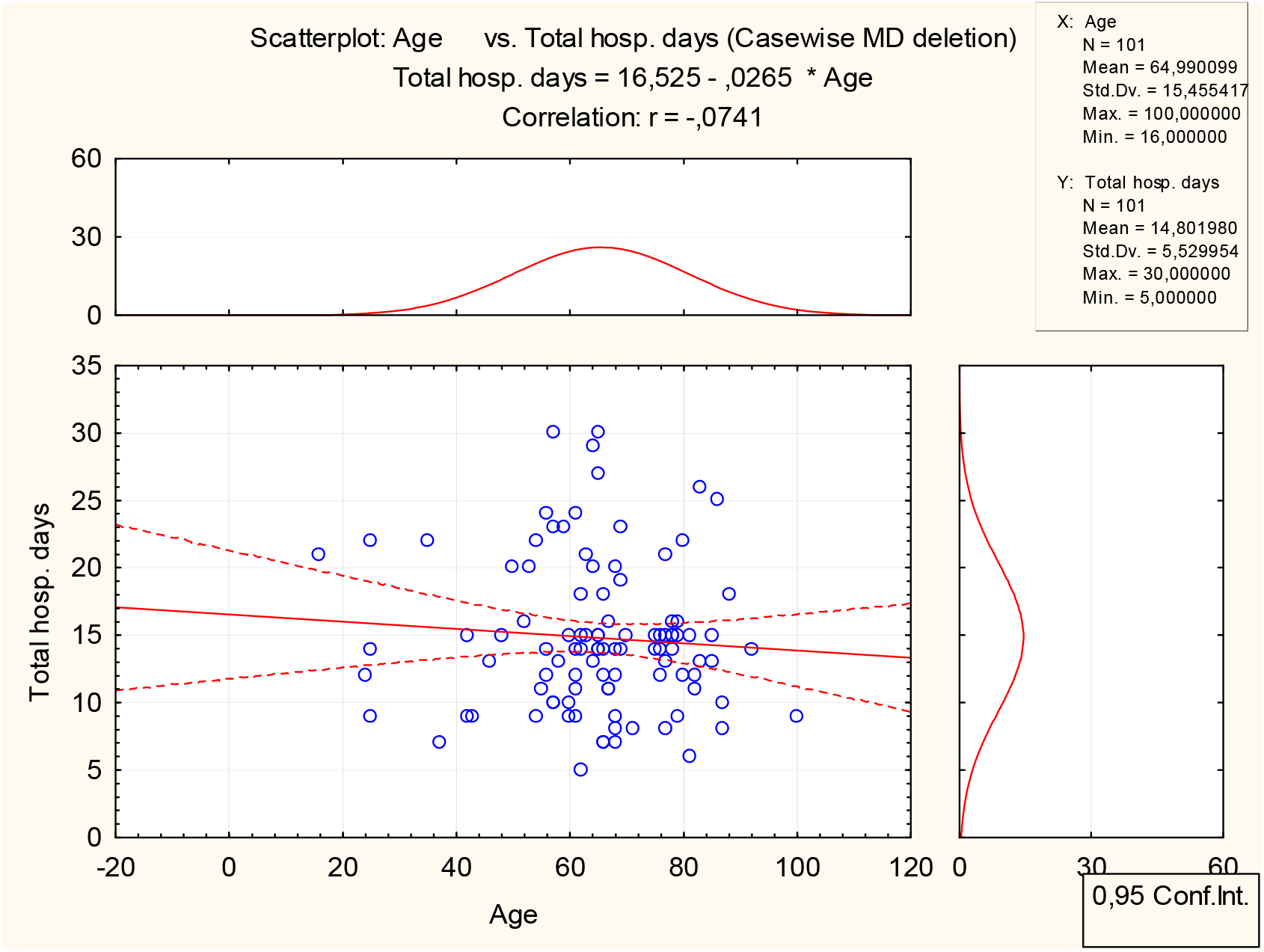
Direct association between age and total hospitalization days. (In the whole sample)

The mean level of the direct bilirubin in the patients living in the city is 45.2872 and in the village is 75.5455, which is statistically significant at t value -2.09959, p< 0.038596. The mean value of the RBC1 in the patients who had ringer and platyphylline is 4.0904 and who didn’t got Ringer and Platyphylline is 2.8000, which is statistically significant at t value 2.16102, p< 0.035935. The mean value of the Ht2 in the patients who had ringer and platyphylline is 155.9062 and who didn’t got Ringer and Platyphylline is 572.6667, which is statistically significant at t value -3.61719, p< 0.000583.

The mean value of the RBC1 in the patients who had papaverine is 4.0904 and who didn’t papaverine is 2.8000, which is statistically significant at t value 2.16102, p< 0. 035935. The mean value of the AST2 in the patients who had papaverine is 152.2208 and who didn’t get papaverine is 389.4286, which is statistically significant at t value -2.62447, p< 0.010349. The mean value of total bilirubin 1 in patients who had NaCl 0.9%/metamizole/diphenhydramine (DPH, Dimedrolum) is 95.9120 and those who did not get it is 222.8938, which is statistically significant at the t value -2.43377, p< 0.016941. The mean value of Hb2 in the patients who had NaCl 0.9% and aprotinin is 20.1080 and who did not have NaCl 0.9% and aprotinin is 57.4695, which is statistically significant at the value of t -3.00739, p< 0.003498.

The mean value of the WBC2 in the patients who had NaCl 0.9% and aprotinin is 4.4547and who didn’t get NaCl 0.9% and aprotinin is 6.1635, which is statistically significant at t value - 2.90601, p< 0.005442. The mean value of RBC2 in the patients who had NaCl 0.9% and aprotinin is 29.5100 and who did not get NaCl 0.9% and aprotinin is 8.3717, which is statistically significant with t value - 3.70938, p< 0.000875. The mean value of the ALT2 in the patients who had NaCl 0.9% and aprotinin is 132.3320 and who didn’t get NaCl 0.9% and aprotinin is 89.2200, which is statistically significant at t value 2.43160, p< 0.017059.

The mean value of the indirect bilirubin 2 in the patients who had NaCl 0.9% and aprotinin is 7.1330 and who didn’t get NaCl 0.9% and aprotinin is 23.6954, which is statistically significant at t value -2.25719, p< 0.026416. The mean age of the patients who had 5% glucose solution / potassium chloride / manganese sulfate is 67.7538 and who didn’t get glucose solution 5%/ potassium chloride/manganese sulfate is 60.0000, which is statistically significant at the t value - 2.47599, p< 0.014984. The mean Hb2 in the patients who had glucose solution 5%/potassium chloride/manganese sulfate is 31.6400 and who didn’t get glucose solution 5%/potassium chloride/manganese sulfate is 74.2483, which is statistically significant at t value 3.64924, p< 0.000461.

The mean WBC1 in the patients who had glucose solution 5%/potassium chloride /manganese sulfate is 7.6327 and who didn’t get glucose solution 5%/potassium chloride/manganese sulfate is 9.2879, which is statistically significant at t value 2.21303, p< 0.029782. The mean age of the patients who had pancreatin is 56.1500 and who didn’t get pancreatin is 67.1728, which is statistically significant at t value 2.96557, p< 0.003786

In terms of dependent values of the Hb1/Hb2, WBC1/WBC2, RBC1/RBC2, Ht1/Ht2, AST1/AST2, total bilirubin 1/total bilirubin 2, direct bilirubin 1/ direct bilirubin 2, and indirect bilirubin 1/ indirect bilirubin 2, are dependent and related values from the first and second analysis except in ALT1/ALT2, there is no statistically confirmed dependence, t value for the dependent parameters, t value 12.05875; p<0.000000, t value 6.01233; p< 0.000001, t value - 2.93772; p< 0.007192, t value -2.17036; p< 0.049094, t value -2.85976; p< 0.005647, t value - 2.15884; p< 0.033681, t value 2.12973; p< 0.036230, t value 5.48481; p< 0.000000, respectively.

By the etiology of cholithiasis, the frequency of gallstones is reported in 37 (36.63366%) patients, acute pancreatitis in 23 (22.77228%) patients, post-cholecystectomy syndrome (PCS) in 8 (7.92079 %) patients, Hilar cholangiocarcinoma (HC, Klatskin tumor) in 1 (0.99010 %) patient, pancreatic cancer in 12 (11.88119 %) patients, acute biliary pancreatitis in 8 (7.92079 %) patients, pancreatic pseudotumor in 4 (3.96040%) patients, acute cholecystitis in 3 (2.97030 %) patients, papillary tumor in 2 (1.98020 %) patients, and pancreatic cyst in 3 (2.97030%) patients. The frequency of patients who received Ringer and Platyphylline is 98 (97.02970 %) patients and 3 (2.97030%) did not receive it. The frequency of patients who received papaverine is 94 (93.06931%) patients and 7 (6.93069%) did not receive it. The frequency of patients who received NaCl 0.9% / metamizole / diphenhydramine (DPH, Dimedrolum) is 84 (83.16832 %) patients and 17 (16.83168%) did not receive it. The frequency of patients who received 0.9% NaCl / aprotinin (Aprotex) is 28 (27.72277 %) patients and 73 (72.27723%) did not receive it. The frequency of patients who required glucose solution 5%/potassium chloride/manganese sulfate is 36 (35.64356 %) patients and 65 (64.35644 %) didn’t receive it. The frequency of patients who received Omeprazole is 46 (45.54455%) patients and 55 (54.45545%) did not receive it. The frequency of patients who received pancreatin is 20 (19.80198 %) patients and 81 (80.19802%) did not receive it. The frequency of patients who got Gordox is 101 (100.0000 %) patients and 0 (0.0000 %) didn’t receive it.

The frequency of patients who underwent EPST surgery is 10 (9.90099 %) patients, CBD surgery 6 (5.94059 %) patients, CDBD surgery 1 (0.99010%) patient, cholecystostomy surgery 1 (0.99010 %) patients, hepaticocholecystoenterostomy surgery 2 (1.98020 %) patients, conservative treatment in 81 (80.19802 %) patients.

The frequency of patients who received antibacterial therapy is 70 (69.30693%) patients and 31 (30.69307%) did not receive it. Out of 101, 25 (24.75248%) patients received Ciprofloxacin, 7 (6.93069%) received Cefazolin, 14 (13.86139 %) received Ceftriaxone, 9 (8.91089 %) received Ciprofloxacin and Metronidazole, 1 (0.99010 %) received Ciprofloxacin/Metronidazole/Ceftriaxone, 1 (0.99010%) received Metronidazole and Ceftriaxone, 2 (1.98020 %) received Metronidazole, 3 (2.97030 %) received Ceftriaxone and Metronidazole, 1 (0.99010 %) received Ciprofloxacin/Ceftriaxone/Metronidazole, 2 (1.98020 %) received Ciprofloxacin and Ceftriaxone, 2 (1.98020 %) received Ceftazidime, 1 (s0.99010 %) received Ceftriaxone and Amikacin, 1 (0.99010 %) received Ciprofloxacin and Metronidazole, and 1 (0.99010 %) received Ceftriaxone and Ceftazidime.

The mean total hospitalization days of patients with cholithiasis due to gallstones is 15.32432 days (Std Err 1.014944). The mean total hospitalization days of patients with cholithiasis due to Acute pancreatitis is 15.60870 days (Std Err 1.104678). The mean total hospitalization days of patients with cholithiasis due to post-cholecystectomy syndrome (PCS) is 15.75000 days (Std Err 0.000). The mean total hospitalization days of patients with cholithiasis due to Hilar cholangiocarcinoma (HC, Klatskin tumor) is 9.00000 days (Std Err 0.000). The mean total hospitalization days of patients with cholithiasis due to pancreatic cancer is 12.16667 days (Std Err 1.021239). The mean total hospitalization days of patients with cholithiasis due to acute biliary pancreatitis is 13.00000 days (Std Err 1.982062). The mean total hospitalization days of patients with cholithiasis due to pancreatic pseudotumor is 16.00000 days (Std Err 4.564355). The mean total hospitalization days of patients with cholithiasis due to acute cholecystitis is 18.00000 days (Std Err 3.000000). The mean total hospitalization days of patients with cholithiasis due to papillary tumor is 14.00000 days (Std Err 0.000000). The mean total hospitalization days of patients with cholithiasis due to pancreatic cyst is 12.66667 days (Std Err 4.702245).

Of 101, 31 (30.69%) did not receive antibacterial therapy and 70 (69.31%) patients received antibacterial therapy. The mean total hospitalization days of the patients who received Ciprofloxacin 14.84000 days (Std Err 0.926787), Cefazolin 15.57143 days (Std Err 2.213133) Ceftriaxone 15.28571(Std Err 1.948312) days, Ciprofloxacin and Metronidazole 13.88889 days (Std Err 2.130757), Ciprofloxacin/Metronidazole/Ceftriaxone 29.00000 days, Metronidazole and Ceftriaxone 27.00000 days, Metronidazole 14.00000 days (Std Err 1.000000), Ciprofloxacin and Ceftriaxone 11.50000 days (Std Err 1.333333), Ceftazidime 16.00000 days (Std Err 4.000000), and Ceftriaxone with Amikacin 25.00000 days.

Of 101 patients, 20 (19.80%) patients underwent surgical treatment and 81 (80.20%) patients did not require surgery. The mean total hospitalization days for patients who passed EPST surgery is 16.20000 days (Std Err 1.008850), CBD 21.50000 days (Std Err 1.565248), CDBD 25.00000 days, cholecystostomy 14.00000 days, and hepaticocholecystoenterostomy 16.00000 days (Std Err 2.000000).

The mean age of patients with gallstones 66.4054 years (Std Err 2.45667), acute pancreatitis 59.0000 years (Std Err 3.71728), post-cholecystectomy syndrome (PCS) 72.8750 years (Std Err 2.25545), Hilar cholangiocarcinoma (HC, Klatskin tumor) 100.0000 years, pancreatic cancer 72.4167 years (Std Err 2.49381), acute biliary pancreatitis 63.2500 years (Std Err 4.07847), pancreatic pseudotumor 67.7500 years (Std Err 3.52077), acute cholecystitis 59.3333 years (Std Err 1.76383), papillary tumor 63.5000 years (Std Err 2.50000), pancreatic cyst 38.6667 years (Std Err 13.66667). The age and etiology of cholithiasis are statistically correlated with each other, as well as the total hospitalization days and the type of surgery. (*Figure 2,3*)

**Figure 2:**
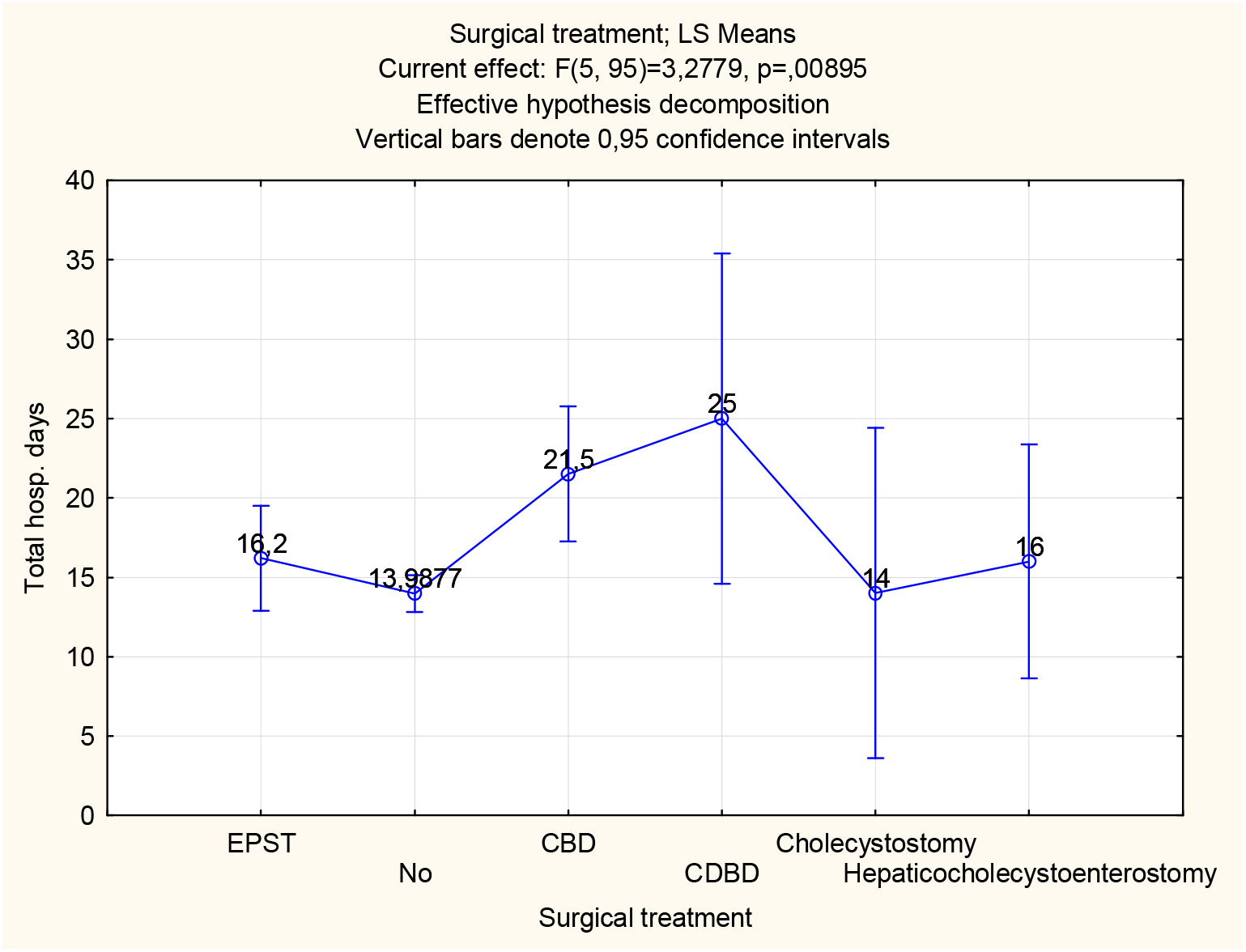
Total hospitalization days is associated with the type of surgery. (In the whole sample)

**Figure 3:**
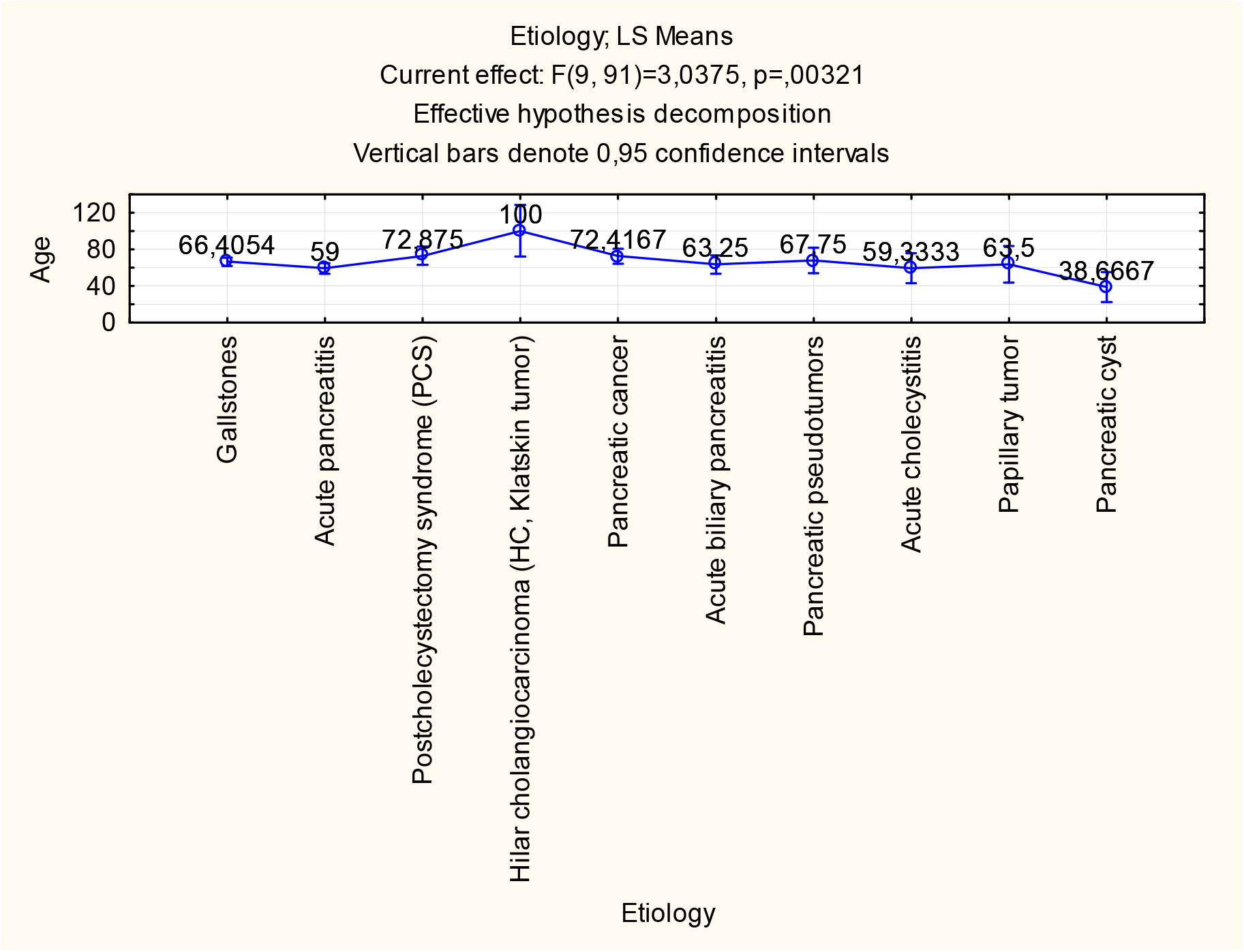
Association between age and the etiology of cholithiasis. (In the whole sample)

In the male group, the mean age is 62.7805 (min; max, 16.00000; 86.000) years, (median; Std Err, 66.0000; 2.40541). In the female group, the mean age is 66.5000 (min; max, 24.00000; 100.000) years, (median; Std Err, 65.5000; 1.99300). In male group, the mean total hospitalization days 15.8537 (min; max, 5.00000; 30.000) days, (median; Std Err, 15.0000; 0.89071). In the female group, the mean total hospitalization days 14.0833 (min; max, 6.00000; 29.000) days, (median; Std Err, 13.5000; 0.68901). In male group, the mean Hb1 is 128.9677 (min; max, 96.00000; 170.000), (median; Std Err, 124.0000; 3.32843). In the female group, the mean Hb1 is 122.3261 (min; max, 80.00000; 160.000), (median; Std Err, 125.0000; 2.71266). In male group, the mean Hb2 is 46.4194 (min; max, 4.00000; 156.000), (median; Std Err, 10.9000; 10.07661). In the female group, the mean Hb2 is 46.3094 (min; max, 5.10000; 152.000), (median; Std Err, 12.1000; 7.43395). In male group, the mean WBC1 is 8.5667 (min; max, 3.40000; 16.500), (median; Std Err, 7.4000; 0.59239). In the female group, the mean WBC1 is 7.9906 (min; max, 3.70000; 3.70000), (median; Std Err, 7.0500; 0.46924). In male group, the mean WBC2 is 5.6535 (min; max, 3.40000; 10.200), (median; Std Err, 4.9500; 0.41202). In the female group, the mean WBC2 is 5.6813 (min; max, 3.34000; 14.700), (median; Std Err, 5.0000; 0.39039). In male group, the mean RBC1 is 4.06944444 (min; max, 2.90000; 5.170), (median; Std Err, 4.1050; 0.16427). In the female group, the mean RBC1 is 4.0600 (min; max, 2.80000; 4.800), (median; Std Err, 4.1150; 0.10413). In male group, the mean RBC2 is 13.5627273 (min; max, 3.30000; 42.000), (median; Std Err, 3.7500; 5.07498). In the female group, the mean RBC2 is 12.9150 (min; max, 3.20000; 44.000), (median; Std Err, 4.4300; 3.51281). In the male group, the mean Ht1 is 35.6666667 (min; max, 27.00000; 43.000), (median; Std Err, 35.0000; 2.47207). In the female group, the mean Ht1 is 32.9619 (min; max, 0.39000; 42.000), (median; Std Err, 36.5000; 2.40433). In male group, the mean Ht2 is 177.076923 (min; max, 22.00000; 1500.000), (median; Std Err, 97.0000; 57.21979). In the female group, the mean Ht2 is 172.9756 (min; max, 29.00000; 491.000), (median; Std Err, 133.0000; 22.65856). In the male group, the mean AST1 is 94.8129032 (min; max, 14.00000; 278.000), (median; Std Err, 88.0000; 11.45956). In the female group, the mean AST1 is 100.8163 (min; max, 12.00000; 568.000), (median; Std Err, 74.0000; 14.12305). In male group, the mean AST2 is 171.666667 (min; max, 14.00000; 1900.000), (median; Std Err, 97.0000; 53.38399). In the female group, the mean AST2 is 172.2292 (min; max, 4.00000; 598.000), (median; Std Err, 104.5000; 21.76342). In male group, the mean ALT1 is 120.248387 (min; max, 20.00000; 373.000), (median; Std Err, 108.7000; 13.97935). In the female group, the mean ALT1 is 126.5102 (min; max, 16.00000; 746.000), (median; Std Err, 90.0000; 18.11587). In male group, the mean ALT2 is 103.368421 (min; max, 18.00000; 427.000), (median; Std Err, 85.0000; 13.20499). In the female group, the mean ALT2 is 99.6077 (min; max, 10.00000; 416.000), (median; Std Err, 76.0000; 10.41279). In the male group, the mean total bilirubin 1 is 121.394444 (min; max, 5.40000; 512.000), (median; Std Err, 81.0000; 21.12038). In the female group, the mean total bilirubin 1 is 116.1727 (min; max, 5.40000; 1645.000), (median; Std Err, 49.0000; 30.94943). In male group, the mean total bilirubin 2 is 71.0794872 (min; max, 2.70000; 272.000), (median; Std Err, 60.0000; 9.21227). In the female group, the mean total bilirubin 2 is 73.6088 (min; max, 0.00000; 451.000), (median; Std Err, 42.0000; 11.30112). In the male group, the mean direct bilirubin 1 is 71.8055556 (min; max, 0.00000; 302.000), (median; Std Err, 37.3000; 12.84791). In the female group, the mean direct bilirubin 1 is 52.1364 (min; max, 0.00000; 214.000), (median; Std Err, 18.0000; 8.69360). In male group, the mean direct bilirubin 2 is 48.0648649 (min; max, 0.00000; 155.000), (median; Std Err, 47.0000; 5.83317). In the female group, the mean direct bilirubin 2 is 42.3846 (min; max, 0.00000; 231.000), (median; Std Err, 26.0000; 6.48745). In male group, the mean indirect bilirubin 1 is 50.8944444 (min; max, 0.00000; 210.000), (median; Std Err, 28.4000; 9.27019). In the female group, the mean indirect bilirubin 1 is 45.9509 (min; max, 0.00000; 432.000), (median; Std Err, 25.0000; 8.80246).

In male group, the mean level of indirect bilirubin 2 is 16.6358974 (min; max, 0.00000; 104.000), (median; Std Err, 6.6000; 3.73463). In the female group, the mean level of indirect bilirubin 2 is 20.4526 (min; max, 4.00000; 220.000), (median; Std Err, 5.9000; 5.27445). In the male group, the mean glucose level is 6.097 (min; max, 4.30000; 7.800), (median; Std Err, 6.2000; 0.42751). In the female group, the mean glucose level is 5.9221 (min; max, 4.00000; 13.380), (median; Std Err, 5.2400; 0.52191).

In a male group, the frequency of patients who live in the city is 19 (46.34146%) patients and 22 (53.65854%) patients’ lives in the village. Whereas, in female group, the frequency of patients who lives in the city is 35 (58.33333%) patients and 25 (41.66667%) patients’ lives in the village. In male group, the frequency of cholthiasis due to gallstones is 11 (26.82927%), acute cholecystitis 1 (1.66667 %), acute pancreatitis 11 (26.82927 %), pancreatic pseudotumor 3 (7.31707%), pancreatic cyst 1 (1.66667 %), post-cholecystectomy syndrome (PCS) 2 (4.87805 %), papillary tumor 1 (1.66667 %), pancreatic cancer 7 (17.07317%), and acute biliary pancreatitis 2 (4.87805 %). Whereas, in the female group, the frequency of cholthiasis due to gallstones is 26 (43.33333%), acute cholecystitis 2 (4.87805 %), acute pancreatitis 12 (20.00000 %), pancreatic pseudotumor 1 (1.66667 %), pancreatic cyst 2 (4.87805 %), post-cholecystectomy syndrome (PCS) 6 (10.00000 %), papillary tumor 1 (2.43902 %), pancreatic cancer 5 (8.33333 %), acute biliary pancreatitis 6 (10.00000 %), and Hilar cholangiocarcinoma (HC, Klatskin tumor) 1 (1.66667 %). In male group, the frequency of patients who received Ringer’s solution and Platyphylline hydrotartrate is 39 (95.12195 %) patients and 2 (4.87805%) patients didn’t receive it. Whereas, in the female group, the frequency of patients who received Ringer ‘s solution and Platyphylline hydrotartrate is 59 (98.33333%) patients and 1 (1.66667%) patient did not. In male group, the frequency of patients who received papaverine hydrochloride is 35 (85.36585%) patients and 6 (14.63415%) patients didn’t receive it. Whereas, in the female group, the frequency of patients who received papaverine hydrochloride is 59 (98.33333 %) patients and 1 (1.66667 %) patient did not receive it. In male group, the frequency of patients who received NaCl 0.9%/metamizole/diphenhydramine is 33 (80.48780 %) patients and 8 (19.51220 %) patients didn’t receive it. Whereas, in the female group, the frequency of patients who received NaCl 0.9%/metamizole/diphenhydramine is 51 (85.00000 %) patients and 9 (15.00000 %) patients didn’t receive it. In male group, the frequency of patients who received 0.9% NaCl and aprotinin (Aprotex) is 8 (19.51220 %) patients and 33 (80.48780 %) patients did not receive it. While, in the female group, the frequency of patients who received 0.9% NaCl 0.9% and aprotinin (Aprotex) is 20 (33.33333 %) patients and 40 (66.66667 %) patients did not receive it. In male group, the frequency of patients who received 5% glucose solution / potassium chloride / manganese sulfate is 26 (63.41463 %) patients and 15 (36.58537 %) patients did not receive it. Whereas, in the female group, the frequency of patients who received a 5% glucose solution / potassium chloride / manganese sulfate is 39 (65.00000 %) patients and 21 (35.00000%) patients did not receive it.

In the male group, 31 (75.61%%) patients received antibiotics and 10 (24.39024%) did not receive antibiotics. In the male group, the frequency of patients who received Ciprofloxacin is 10 (24.39024%), Cefazolin 6 (14.63415%), Ceftriaxone 5 (12.19512%), Ciprofloxacin and Metronidazole 6 (14.63415%), Ceftriaxone and Amikacin 1 (2.43902%), Ceftriaxone and Metronidazole 1 (2.43902%), Ceftriaxone and Ceftazidime 1 (2.43902%), Ciprofloxacin and Ceftriaxone 1 (2.43902%), and Ceftazidime 2 (3.33333 %). However, in the female group, 39 (65.00%) patients received antibiotics and 21 (35.00000%) did not receive antibiotics. In the female group, the frequency of patients who received Ciprofloxacin is 15 (25.00000%), Cefazolin 1 (1.66667 %), Ceftriaxone 9 (15.00000 %), Ciprofloxacin and Metronidazole 4 (6.66667 %), Ciprofloxacin/Metronidazole/Ceftriaxone 2 (3.33333 %), Ceftriaxone and Metronidazole 3 (5.00000 %), Metronidazole 2 (3.33333%), Ciprofloxacin and Ceftriaxone 1 (1.66667 %), and Ceftazidime 2 (3.33333 %).

In male group, the frequency of patients who received Omeprazole is 18 (43.90244 %) patients and 23 (56.09756 %) patients didn’t receive it. Whereas in the female group, the frequency of patients who received Omeprazole is 28 (46.66667 %) patients and 32 (53.33333 %) patients did not receive it. In male group, the frequency of patients who received pancreatin is 8 (19.51220%) patients and 33 (80.48780 %) patients didn’t receive it. However, in the female group, the frequency of patients who received pancreatin is 12 (20.00000 %) patients and 48 (80.00000 %) patients who did not receive it.

In the male group, the frequency of patients who received Gordox is 41 (100.0000%) patients and 0 (0.0000 %) patients did not receive it. However, in the female group, the frequency of patients who received Gordox is 60 (100.0000%) patients and 0 (0.0000 %) patients didn’t receive it. In male group, the frequency of patients who underwent surgery is 9 (21.95%) patients and 32 (78.04878 %) patients who did not pass surgical treatment. Of the 9 patients, 3 (7.31707 %) patients underwent CBD surgery, 1 (2.43902%) patient underwent CDBD surgery, 1 (2.43902 %) patient underwent cholecystostomy surgery, 2 (4.87805 %) patients underwent EPST surgery, 2 (4.87805 %) patients underwent hepaticocholecystoenterostomy surgery, while in the female group, the frequency of patients who underwent surgery is 11 (18.33 %) patients and 49 (81.66667%) patients did not undergo surgical treatment. Out of the 11 patients, 3 (5.00000%) patients underwent CBD surgery, 8 (13.33333 %) patients underwent EPST surgery. In terms of gender, in male group, the mean level of total bilirubin 1 in the patients living in the city 47.2600 mmol/L, whereas, in the male patients living in the village, the mean level of total bilirubin 1is 174.3476 mmol/L, which is statistically significant difference at t value -3.37946, p<0.001836. The mean level of total bilirubin 2 in the patients living in the city 48.5235 mmol/L, whereas, in the male patients living in the village, the mean level of total bilirubin 2 is 88.5091 mmol/L, which is statistically significant difference at t value -2.26645, p< 0.029362. The mean level of direct bilirubin 1 in the patients living in the city 25.0800 mmol/L, whereas, in the male patients living in the village, the mean level of direct bilirubin 1 is 105.1810 mmol/L, which is statistically significant difference at t value -3.54552, p<0.001165. The mean level of indirect bilirubin 1 in the patients living in the city 25.3133 mmol/L, whereas, in the male patients living in the village, the mean level of indirect bilirubin 1 is 69.1667 mmol/L, which is statistically significant difference at t value -2.50120, p<0.017357.

In the male group, the mean Ht2 level in the patients who received Ringer ‘s solution and Platyphylline hydrotartrate is 122.0833, while in the male patients who did not receive it, the mean level of Ht2 is 837.000, which is a statistically significant difference at t value -4.37231, p< 0.000205. In the female group, the mean level of Hb1 in the patients who got Ringer ‘s solution and Platyphylline hydrotartrate is 123.2667, whereas, in the female patients who did not receive it, the mean level of Hb1 is 80.0000, which is statistically significant difference at t value 2.45212, p< 0.018239. In the female group, the mean level of RBC1 in the patients who got Ringer ‘s solution and Platyphylline hydrotartrate is 4.1034, whereas, in the female patients who did not receive it, the mean level of RBC1 is 2.8000, which is statistically significant difference at t value 2.42954, p< 0.021785. In male group, the mean level of AST2 in the patients who received Ringer solution and Platyphylline hydrotartrate is 122.1765, whereas, in the male patients who did not receive it, the mean level of AST2 is 1013.000, which is a statistically significant difference at the t value -4.93593, p< 0.000021.

In the male group, the mean level of AST1 in the patients who got papaverine hydrochloride is 85.9704, whereas, in the male patients who did not receive it, the mean level of AST1 is 154.5000, which is statistically significant difference at t value -2.11803, p< 0.042865. In the female group, the mean level of Hb1 in the patients who got papaverine hydrochloride is 123.2667. whereas, in the female patients who did not receive it, the mean level of Hb1 is 80.0000, which is statistically significant difference at t value 2.45212, p< 0.018239. In the female group, the mean level of RBC1 in the patients who got papaverine hydrochloride is 4.1034, whereas, in the female patients who did not receive it, the mean level of RBC1 is 2.8000, which is statistically significant difference at t value 2.42954, p< 0.021785. In the male group, the mean level of AST2 in patients who got papaverine hydrochloride is 117.8667, whereas, in the male patients who did not receive it, the mean level of AST2 is a 440.6667, which is statistically significant difference at t value -2.40216, p< 0.021907. In male group, the mean level of ALT1 in the patients who got papaverine hydrochloride is 106.9519, while, in the male patients who did not receive it, the mean level of ALT1 is a 210.0000, which is statistically significant difference at t value -2.72248, p< 0.010850.

In the male group, the mean level of Ht2 in the patients who got NaCl 0.9%/metamizole/diphenhydramine (DPH, Dimedrolum) is 122.0952, whereas, in the male patients who did not receive it, the mean level of Ht2 is 408.0000, which is statistically significant difference at t value -2.09910, p< 0.046508. In the female group, the mean level of Hb1 in the patients who got NaCl 0.9%/metamizole/diphenhydramine (DPH, Dimedrolum) is 124.5897, whereas, in the female patients who did not receive, the mean level of Hb1 is 109.7143, which is statistically significant difference at t value 2.03750, p< 0.047642. In male group, the mean AST1 level in the patients who got NaCl 0.9%/ metamizole / diphenhydramine (DPH, Dimedrolum) is 77.2480, while, in the male patients who did not receive, the mean level of AST1 is 168.0000, which is a statistically significant difference at t value -3.74782, p< 0.000790. In male group, the mean level of ALT1 in the patients who got NaCl 0.9%/metamizole/diphenhydramine (DPH, Dimedrolum) is 102.4280, whereas, in the male patients who did not receive, the mean level of ALT1 is 194.5000, which is statistically significant difference at t value -2.90742, p< 0.006919. In the female group, the mean level of total bilirubin 1 in patients who received NaCl 0.9%/ metamizole / diphenhydramine (DPH, Dimedrolum) is 84.2652, while in women who did not receive it, the mean level of total bilirubin 1 is 279.2556, which is statistically significant difference with t value -2.43480, p< 0.018300. In the female group, the mean level of indirect bilirubin 1 in the patients who received 0.9% NaCl 0.9%/metamizole/diphenhydramine (DPH, Dimedrolum) is 37.4435, whereas, in the female patients who did not receive it, the mean level of indirect bilirubin 1 is 89.4333, which is statistically significant difference at t value -2.26722, p< 0.027481.

In male group, the mean level of Hb2 in the patients who got NaCl 0.9% and aprotinin (Aprotex) is 7.4625, whereas, in the male patients who did not receive it, the mean level of Hb2 is 59.9696, which is statistically significant difference at t value - 2.46555, p< 0.019839. In male group, the mean level of RBC2 in patients who got NaCl 0.9% and aprotinin (Aprotex) is 39.0000, whereas, in the male patients who did not receive it, the mean level of RBC2 is 7.9100, which is statistically significant difference at t value -3.37274, p< 0.008221. In the female group, the mean level of RBC2 in the patients who received 0.9% NaCl 0.9% and aprotinin (Aprotex) is 25.7140, whereas, in the female patients who did not receive it, the mean level of RBC2 is 8.6487, which is statistically significant difference at t value 2.33772, p< 0.031150. In the female group, the mean level of WBC2 in the patients who got NaCl 0.9% and aprotinin (Aprotex) is 4.5320, whereas, in the female patients who did not receive, the mean level of WBC2 is 6.2036, which is statistically significant difference at t value -2.08978, p< 0.045217. In male group, the mean level of ALT2 in the patients who got NaCl 0.9% and aprotinin (Aprotex) is 194.3750, whereas, in the male patients who did not receive it, the mean level of ALT2 is 79.1000, which is statistically significant difference at t value -4.32876, p< 0.000114. In male group, the mean level of total bilirubin 2 in the patients who got NaCl 0.9% and aprotinin (Aprotex) is 112.7500, whereas, in the male patients who did not receive it, the mean level of total bilirubin 2 is 60.3258, which is statistically significant difference at t value -2.44356, p< 0.019433. In male group, the mean level of direct bilirubin 2 in the patients who got NaCl 0.9% and aprotinin (Aprotex) is 81.6250, whereas, in the male patients who did not receive it, the mean level of direct bilirubin 2 is 38.8069, which is statistically significant difference at t value -3.44886, p< 0.001484.

In the female group, the mean age of the patients who received the 5% glucose solution / potassium chloride / manganese sulfate is 69.5128 years, while the female patients who did not receive it, the mean age is 60.9048 years, which is statistically significant difference with the value of t -2.12025, p< 0.038273. In male group, the mean Hb2 level in the patients who got glucose solution / potassium chloride / manganese sulfate is 24.4050, whereas, in the male patients who did not receive it, the mean level of Hb2 is 86.4455, which is a statistically significant difference at t value 3.43554, p< 0.001805. In the female group, the mean Hb2 level in patients who received a glucose solution / potassium chloride / manganese sulfate is 35.7743, while in women who did not receive it, the mean Hb2 level of Hb2 is a 66.7944, which is statistically significant difference at the t value 2.03496, p< 0.047069. In male group, the mean level of WBC1 in the patients who got glucose solution 5%/potassium chloride/manganese sulfate is 7.2200, whereas, in the male patients who did not receive it, the mean level of WBC1 is 10.6385, which is statistically significant difference at t value 3.20130, p< 0.003154. In male group, the mean level of WBC2 in the patients who received a 5% glucose solution / potassium chloride / manganese sulfate is 4.8882, while, in the male patients who did not receive it, the mean level of WBC2 is a 6.5889, which is statistically significant difference with the value t 2.26599, p< 0.036018. In male group, the mean level of RBC2 in the patients who got glucose solution 5%/potassium chloride/manganese sulfate is 25.1980, whereas, in the male patients who did not receive, the mean level of RBC2 is 3.8667, which is statistically significant difference at t value -2.64860, p< 0.026537.

In the male group, the mean age of patients who got pancreatin is 50.6250 years, while, in the male patients who did not receive it, the mean age is a 65.7273, which is statistically significant difference at the t value 2.67231, p< 0.010938. In male group, the mean level of glucose in the patients who got pancreatin is 7.4667, whereas, in the male patients who did not receive, the mean level of glucose is 5.5100, which is statistically significant difference at t value -2.76569, p< 0.024459. In the female group, the mean level of AST2 in patients who received pancreatin is 256.2000, while, in the female patients who did not receive it, the mean level of AST2 is 150.1316, which is statistically significant difference at the value of t -2.04521, p< 0.046579.

In terms of dependent samples, in the male group, the mean value of Hb1 is 128.4091 and the mean value of Hb2 is 47.4136, which is statistically significant different at the t value 7.21291, p< 0.000000. However, in the female group, the mean value of Hb1 is 121.0732 and the mean Hb2 is 45.4951, which is statistically significant different with a value of 9.55837, p< 0.000000. In a male group, the mean value of WBC1 is 8.5357 and the mean WBC2 is 5.7571, which is statistically significant different with the value of t 3.49483, p< 0.003953. Whereas, in the female group, the mean value of WBC1 is 8.6240 and the mean value of WBC2 is 5.3836, which is statistically significant different with t value 4.82508, p< 0.000065. In the female group, the mean value of RBC1 is 4.1606 and the mean RBC2 is 12.2053, which is statistically significant different at the t value -2.16063, p< 0.046236. In the female group, the mean value of AST1 is 95.9756 and the mean AST2 is 190.0244, which is statistically significant different at t value - 3.71306, p< 0.000624. In male group, the mean value of total bilirubin 1 is 124.0059 and the mean total bilirubin 2 is 73.7735, which is statistically significant different at t value 2.80437, p< 0.008383. In male group, the mean value of direct bilirubin 1 is 76.4844 and the mean direct bilirubin 2 is 49.8437, which is statistically significant different at t value 2.12517, p< 0.041647. In male group, the mean value of indirect bilirubin 1 is 53.0118 and the mean indirect bilirubin 2 is 18.4824, which is statistically significant different at the t value 5.05326, p< 0.000016. Whereas, in the female group, the mean value of indirect bilirubin 1 is 48.0260 and the mean indirect bilirubin 2 is 21.0998, which is statistically significant different with the t value 3.38443, p< 0.001411.

In male group, the mean total hospitalization days of patients who passed EPST surgery is 14.50000 days. In male group, the mean total hospitalization days of patients who passed hepaticocholecystoenterostomy surgery is 16.00000 (Std Err 2.000000) days. In male group, the mean total hospitalization days of patients who passed CDBD surgery is 25.00000 days. In male group, the mean total hospitalization days of patients who passed cholecystostomy surgery is 14.50000 days. Moreover, in male group, the mean total hospitalization days for patients who passed CBD surgery is 19.33333 (Std Err 2.333333) days. Whereas, the mean total hospitalization days of patients who didn’t pass any surgery is 15.37500 (Std Err 1.065165) days. In female group, the mean total hospitalization days of patients who passed EPST surgery is 16.62500 (Std Err 1.223833) days. Furthermore, in the female group, the mean total hospitalization days of patients who underwent CBD surgery is 23.66667 days (Std Err 2.516611). Whereas, the mean total hospitalization days of patients who didn’t pass any surgery is 13.76667 (Std Err 1.053384) days. (*figure 2*)

In the male group, the mean age of patients who passed EPST surgery is 63.00000 (Std Err 3.000000) years. In male group, the age of patients who passed hepaticocholecystoenterostomy surgery is 71.00000 (Std Err 5.000000) years. In male group, the mean age of patients who passed CDBD surgery is 86.00000 years. In male group, the mean age of patients who underwent cholecystostomy surgery is 78.00000 years. Moreover, in male group, the mean age of patients who passed CBD surgery is 61.00000 (Std Err 2.081666) years. However, the mean age of the patients who did not have surgery is 61.21875 (Std Err 2.910193) years. In female group, the mean age of patients who passed EPST surgery is 74.00000 (Std Err 2.326611) years. Furthermore, in the female group, the mean age of the patients who underwent CBD surgery is 69.00000 years (Std Err 7.023769). Whereas, the mean age of patients who did not pass any surgery is 65.43333 (Std Err 2.937543). (*Table 1*) (*Figure 3*)

**Table 1:**
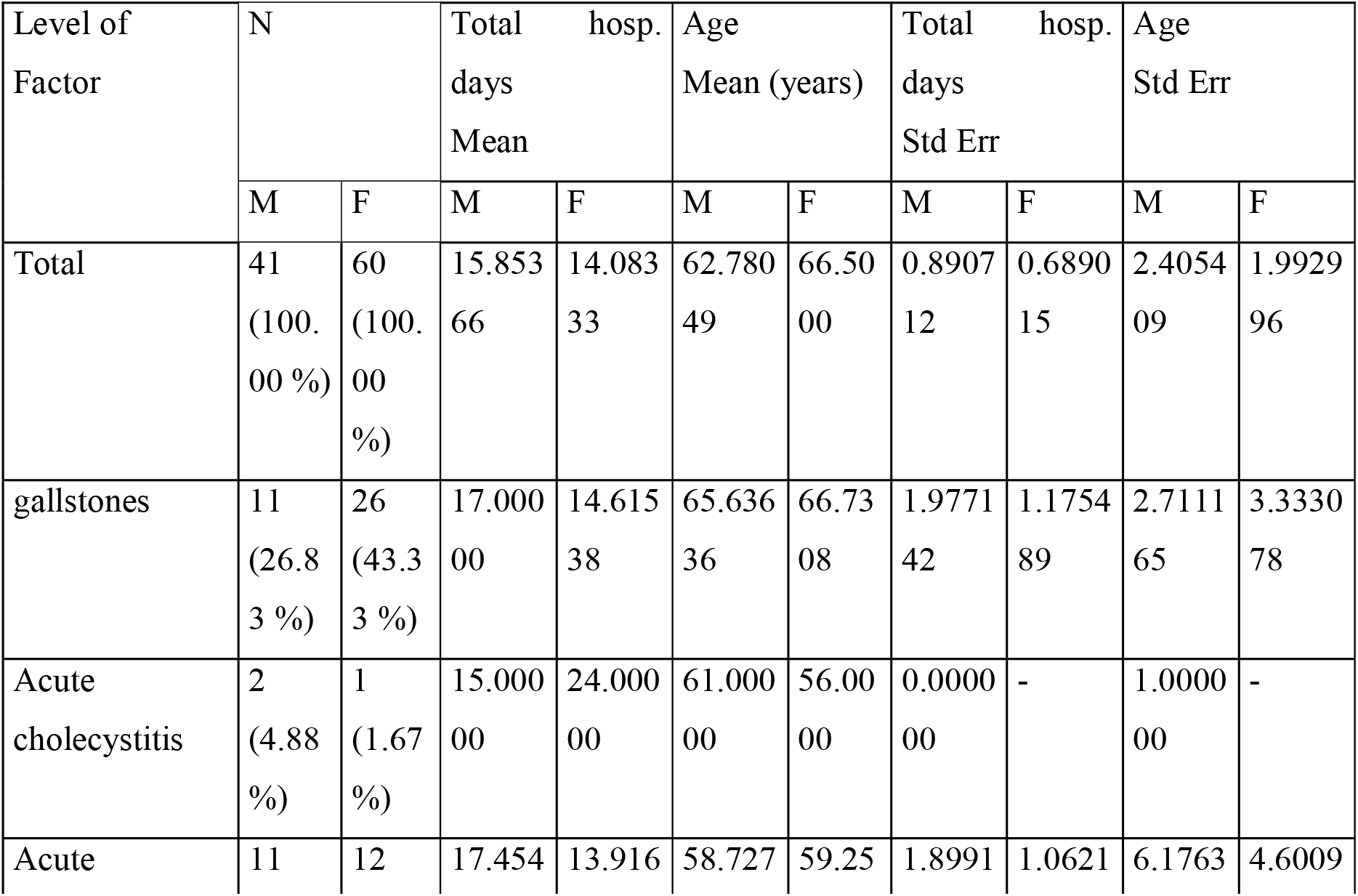
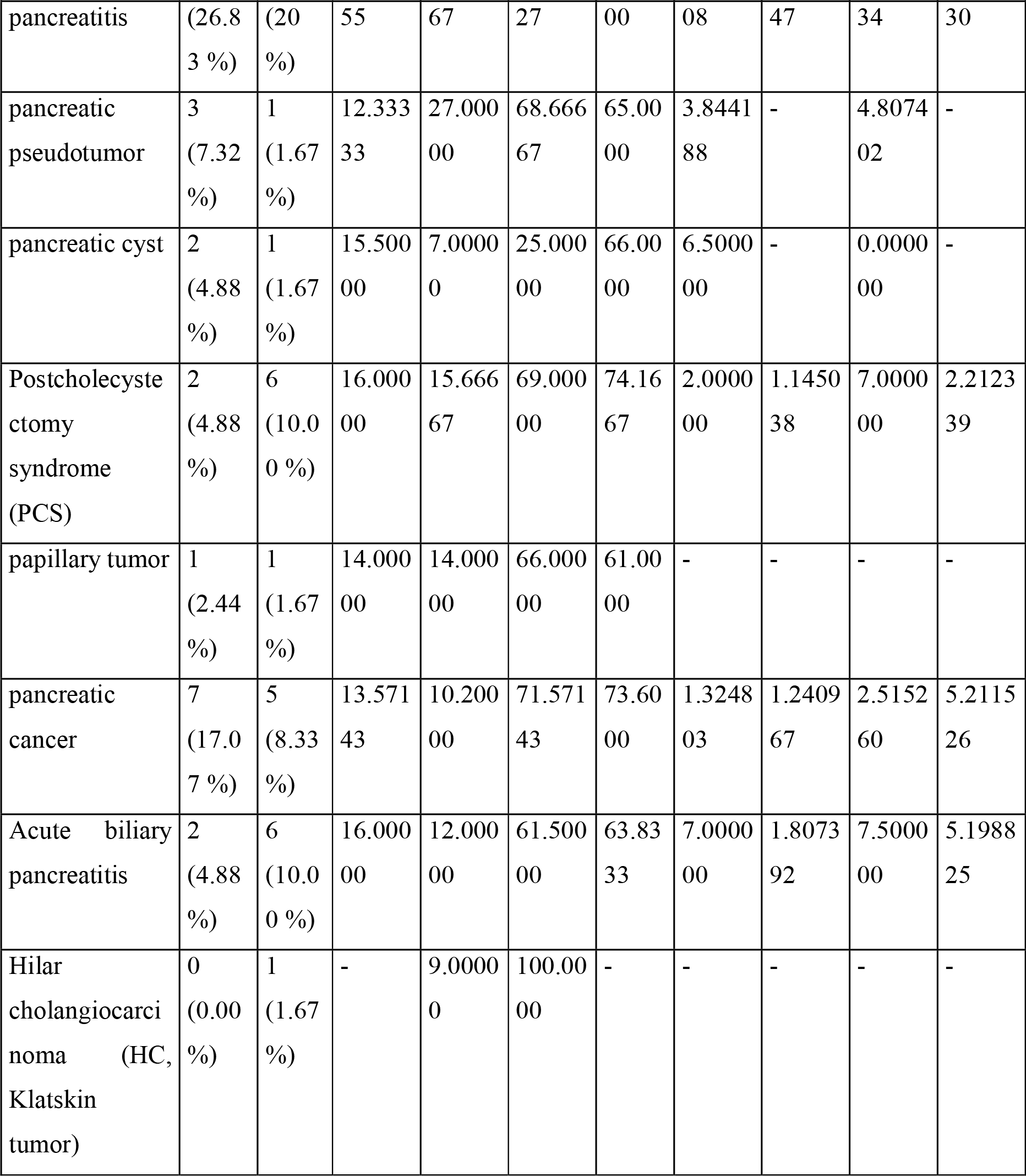
A comparison of the total hospitalization days and the age of the patients in a gender dependent manner.

| Level of Factor | N |  | Total hosp. days Mean |  | Age Mean (years) |  | Total hosp. days Std Err |  | Age Std Err |  |
| --- | --- | --- | --- | --- | --- | --- | --- | --- | --- | --- |
|  | M | F | M | F | M | F | M | F | M | F |
| Total | 41<br>(100.00 %) | 60<br>(100.00 %) | 15.853<br>66 | 14.083<br>33 | 62.780<br>49 | 66.50<br>00 | 0.8907<br>12 | 0.6890<br>15 | 2.4054<br>09 | 1.9929<br>96 |
| gallstones | 11<br>(26.83 %) | 26<br>(43.33 %) | 17.000<br>00 | 14.615<br>38 | 65.636<br>36 | 66.73<br>08 | 1.9771<br>42 | 1.1754<br>89 | 2.7111<br>65 | 3.3330<br>78 |
| Acute cholecystitis | 2<br>(4.88 %) | 1<br>(1.67 %) | 15.000<br>00 | 24.000<br>00 | 61.000<br>00 | 56.00<br>00 | 0.0000<br>00 | - | 1.0000<br>00 | - |
| Acute | 11 | 12 | 17.454 | 13.916 | 58.727 | 59.25 | 1.8991 | 1.0621 | 6.1763 | 4.6009 |

| Level of Factor | N |  | Total hosp. days<br>Mean |  | Age<br>Mean (years) |  | Total hosp. days<br>Std Err |  | Age<br>Std Err |  |
| --- | --- | --- | --- | --- | --- | --- | --- | --- | --- | --- |
|  | M | F | M | F | M | F | M | F | M | F |
| pancreatitis | (26.8<br>3 %) | (20<br>%) | 55 | 67 | 27 | 00 | 08 | 47 | 34 | 30 |
| pancreatic pseudotumor | 3<br>(7.32<br>%) | 1<br>(1.67<br>%) | 12.333<br>33 | 27.000<br>00 | 68.666<br>67 | 65.00<br>00 | 3.8441<br>88 | - | 4.8074<br>02 | - |
| pancreatic cyst | 2<br>(4.88<br>%) | 1<br>(1.67<br>%) | 15.500<br>00 | 7.0000<br>0 | 25.000<br>00 | 66.00<br>00 | 6.5000<br>00 | - | 0.0000<br>00 | - |
| Postcholecystectomy syndrome (PCS) | 2<br>(4.88<br>%) | 6<br>(10.0<br>0 %) | 16.000<br>00 | 15.666<br>67 | 69.000<br>00 | 74.16<br>67 | 2.0000<br>00 | 1.1450<br>38 | 7.0000<br>00 | 2.2123<br>39 |
| papillary tumor | 1<br>(2.44<br>%) | 1<br>(1.67<br>%) | 14.000<br>00 | 14.000<br>00 | 66.000<br>00 | 61.00<br>00 | - | - | - | - |
| pancreatic cancer | 7<br>(17.0<br>7 %) | 5<br>(8.33<br>%) | 13.571<br>43 | 10.200<br>00 | 71.571<br>43 | 73.60<br>00 | 1.3248<br>03 | 1.2409<br>67 | 2.5152<br>60 | 5.2115<br>26 |
| Acute biliary pancreatitis | 2<br>(4.88<br>%) | 6<br>(10.0<br>0 %) | 16.000<br>00 | 12.000<br>00 | 61.500<br>00 | 63.83<br>33 | 7.0000<br>00 | 1.8073<br>92 | 7.5000<br>00 | 5.1988<br>25 |
| Hilar cholangiocarcinoma (HC, Klatskin tumor) | 0<br>(0.00<br>%) | 1<br>(1.67<br>%) | - | 9.0000<br>0 | 100.00<br>00 | - | - | - | - | - |

In the female group, in terms of correlation, a straight association between the age and the Hb1/ Hb2/ RBC2/ Ht1/ AST1, correlation coefficient value -0.999819/-0.999957/0.999847/- 0.999910/0.997818, respectively. In the female group, in terms of correlation, there is a straight association between Hb1 and RBC2/Ht1/AST1, correlation coefficient value - 0.999333/0.999984/-0.998894, respectively. In the female group, in terms of the correlation, a direct association between the total hospitalization days and glucose, correlation coefficient value -1.000000. In the female group, in terms of the correlation, a direct association between the WBC2 and RBC1, correlation coefficient value -0.999857. In the female group, in terms of the correlation, a straight association between the WBC1 and RBC1, correlation coefficient value - 0.998906.

## Discussion

Cholithiasis remains a worldwide issue and cause of obstructive jaundice without reduction in the incidence rate [22, 28–30]. Obstructive jaundice is widely reported in the female group and remains without further and deep elaboration of the underlying causes, pathogenesis, potential risk factors, and preferred treatment methods [31]. Cholithiasis is a crucial risk factor for obstructive jaundice development. However, several other causes can result into obstructive jaundice symptoms including tumors compressing the bile duct. The mechanism of cholithiasis and risk factors for development are various involving genetic predisposition, environmental conditions, and local or and systemic metabolic dysfunction such as dyslipidemia and metabolic syndrome [32]. Recent findings suggested that blood pressure dysregulation such as hypertension involved in the pathogenesis of cholithiasis [33]. The prevenance of cholithiasis in Europe is reported in 20% of the total population [34]. Gallstone has been reported in the ancient populations such as mummified Egyptian priestess [35]. The sample involved patients with a symptom of obstructive jaundice. There are several types of treatment for this pathology. If the cause is in the stone, then they are treated with antispasmodics [36]. If they do not help, then an endoscopic operation with a dissection of the sphincter of Oddi, if it does not help, then choledochotomy. If the cause is a tumor, then this is an operation or palliative treatment, depending on the stage.

Existence of various causes of obstructive jaundice is reported in this study. However, cholithiasis remains the leading cause of obstructive jaundice. The underlying pathophysiological mechanisms of developments of cholithiasis remains poorly understood and further elaboration is required to develop a clinical recommendation on cholithiasis prevention [34]. The treatment plans of cholithiasis in acute symptomatic conditions is indicated for surgical treatment [37–40]. However, in asymptomatic chronic cholithiasis the management plan involved wide spectrum antibiotic, analgesics, and spasmolytics [41, 42]. Therefore, time factor is crucial in term of treatment plan determination [43].

Males are affected by cholithiasis at early age which suggest that young men are probably have specific physiological process that increases the risk of stones formation. Including hormonal changes and metabolic process [44]. Changes in the laboratory parameters are probably related to the gender, living place, treatment choice including surgery or conservative, the used antibiotics, and the

An association between the glucose level and the administration of pancreatin observed in male group. Suggesting a potential regulatory role for glucose or it is regulators, primarily insulin and glucagon, secretion or synthesis in the pancreas. However, previous studies suggested that pancreatin is not involved in the regulation of glucose level and or insulin secretion [45, 46].

In male group, the frequency of patients who underwent surgery is 9 patients and 32 patients who did not pass surgical treatment. Of the 9 patients, 3 patients underwent CBD surgery, 1 patient underwent CDBD surgery, 1 patient underwent cholecystostomy surgery, 2 patients underwent EPST surgery, 2 patients underwent hepaticocholecystoenterostomy surgery, while in the female group, the frequency of patients who underwent surgery is 11 patients and 49 patients did not undergo surgical treatment. Out of the 11 patients, 3 patients underwent CBD surgery, 8 patients underwent EPST surgery. The diversity in the performed types of surgery in male group returns to the clinical presentation and the etiology of obstructive jaundice.

Gallstone disease is associated with increased overall mortality and to death from cardiovascular disease. Gallstones may be considered a possible cardiometabolic risk factor [27, 47]. Operative time and postoperative hospital stay were significantly longer in males than in females [48]. Antibiotic treatment should be started immediately in patients with obstructive, stone-related acute cholangitis. Dissolution of gallstones and symptomatic treatment are also used [34, 43, 49].

## Conclusions

Tendency to early occurrence of obstructive jaundice symptoms in men compared to women. In treatment plans, men and women required the same total hospitalization days. The incidence rate of obstructive jaundice symptoms in females is higher than in males. A straight association between age and the etiology of obstructive jaundice symptoms as well as a straight association between total hospitalization days and the type of surgery.

## Data Availability

All data produced in the present study are available upon reasonable request to the authors

## Declarations

Authors’ contributions: MB analyzed the statistical data, wrote the draft, and revised the final version of the paper, KS collected the data from the hospital. All authors have read and approved the manuscript.

### List of abbreviation

EPST: Endoscopic papillosphincterotomy,
CDBD: Cholecystectomy, Choledochotomy. T-tube drainage of the common bile duct,
CBD: Choledochotomy.
(AST): T-tube drainage of the common bile duct, aspartate transaminase
(ALT): alanine transaminase
RBC: erythrocytes,
WBC: leukocytes,
Ht: hematocrit,
Hb: hemoglobin,
OJ: Obstructive jaundice,
CT: computed tomography

## ETHICS APPROVAL AND CONSENT TO PARTICIPATE

The study approved by the National Research Mordovia State University, Russia, from “Ethics Committee Requirement N8/2 from 30.06.2021”.

## HUMAN AND ANIMAL RIGHTS

No animals were used in this research. All human research procedures followed were in accordance with the ethical standards of the committee responsible for human experimentation (institutional and national), and with the Helsinki Declaration of 1975, as revised in 2013.

## CONSENT FOR PUBLICATION

Written informed consent was obtained from the patients for publication of study results and any accompanying images.

## STANDARDS OF REPORTING

STROBE guideline has been followed.

## AVAILABILITY OF DATA AND MATERIALS

Not applicable.

## ACKNOWLEDGEMENTS

Declared none.

